# Down-regulation of SARS-CoV-2 neutralizing antibodies in vaccinated smokers

**DOI:** 10.1101/2021.12.09.21267349

**Authors:** Jiahui Zhang, Fei Teng, Xiaomei Zhang, Hongye Wang, Te Liang, Shubin Guo, Xiaobo Yu

**Affiliations:** State Key Laboratory of Proteomics, Beijing Proteome Research Center, National Center for Protein Sciences-Beijing (PHOENIX Center), Beijing Institute of Lifeomics, Beijing, 102206, China; Emergency Medicine Clinical Research Center, Beijing Chao-Yang Hospital, Capital Medical University, & Beijing Key Laboratory of Cardiopulmonary Cerebral Resuscitation, Beijing, 100020, China

## Abstract

Vaccination is an effective approach to help control coronavirus disease 2019 (COVID-19). However, since the vaccines produce a heterogenous immune response, the risk of breakthrough infection is increased in vaccinated individuals who generate low levels of neutralizing antibodies (NAbs). It is therefore paramount in the fight against COVID-19 to identify individuals who have a higher risk of breakthrough infection despite being vaccinated. Here we addressed the effect of cigarette smoking on the production of neutralizing antibodies (NAbs) following COVID-19 vaccination since smoking profoundly suppresses the adaptive immune response to pathogen infection and the association between vaccination and smoking remains unclear. The SARS-CoV-2 Spike antibodies and NAbs (days 0, 14, 42, and 90) were measured in 164 participants received two vaccine doses of an inactivated vaccine (Sinovac-CoronaVac) longitudinally. Anti-Spike antibodies was elevated 14 and 42 days after COVID-19 vaccination compared to baseline (i.e., “Day 0”). Notably, RBD antibodies showed significantly higher expression in the nonsmoking group (n=153) than the smoking (n=11) group on day 42 (p<0.0001, Student’s t-test). NAbs continually increased after the first and second vaccine dose, peaking on day 42. NAbs titers then significantly decreased until day 90. Compared to nonsmokers, the NAb levels in smokers remained low throughout the period of testing. The median NAb titers in the smoking group was 1.40-, 1.32-, or 3.00-fold lower than that of nonsmoking group on day 14, 42, or 90, respectively. Altogether, our results indicate that smoking is a specific risk factor for COVID-19 breakthrough infection following vaccination.

## Text

As of December 7, 2021, coronavirus disease 2019 (COVID-19) has infected more than 2.6 billion people and caused over 5 million deaths worldwide. Vaccination is an effective approach to help control COVID-19 with 8.17 billion vaccine doses thus far administered^1^. However, since the vaccines produce a heterogenous immune response, the risk of breakthrough infection is increased in vaccinated individuals who generate low levels of neutralizing antibodies (NAbs)^2^. It is therefore paramount in the fight against COVID-19 to identify individuals who have a higher risk of breakthrough infection despite being vaccinated.

In this study, we addressed the effect of cigarette smoking on the production of NAbs following COVID-19 vaccination since smoking profoundly suppresses the adaptive immune response to pathogen infection^3^,^4^. We recruited 164 participants between 20 and 58 years old from Beijing Chaoyang Hospital from December 2020 to March 2021, who were divided into smoking and nonsmoking groups (Table S1 and Supplementary Methods). The participants received two vaccine doses of an inactivated whole-virion SARS-CoV-2 vaccine (Sinovac-CoronaVac) two weeks apart (i.e., “Day 0” and “Day 14”), and their serum was collected longitudinally on days 0, 14, 42, and 90 (Table S2). Since the SARS-CoV-2 Spike (S) protein is required for viral entry, serological antibodies on days 0, 14, and 42 targeting Spike (S) protein and its specific domains, including subunit 1 (S1), receptor binding domain (RBD), and subunit 2 extracellular domain (S2ECD), were detected using a protein array (Supplementary Methods).

Antibody levels to S protein and domains (S1, S2ECD, RBD) were elevated 14 and 42 days after COVID-19 vaccination compared to baseline in both participant groups (i.e., “Day 0”). Moreover, RBD antibodies showed significantly higher expression in the nonsmoking group (n=153) than the smoking (n=11) group on day 42 (p<0.0001, Student’s *t*-test) (Figure 1A and Figures S1-S4).

**Figure 1:**
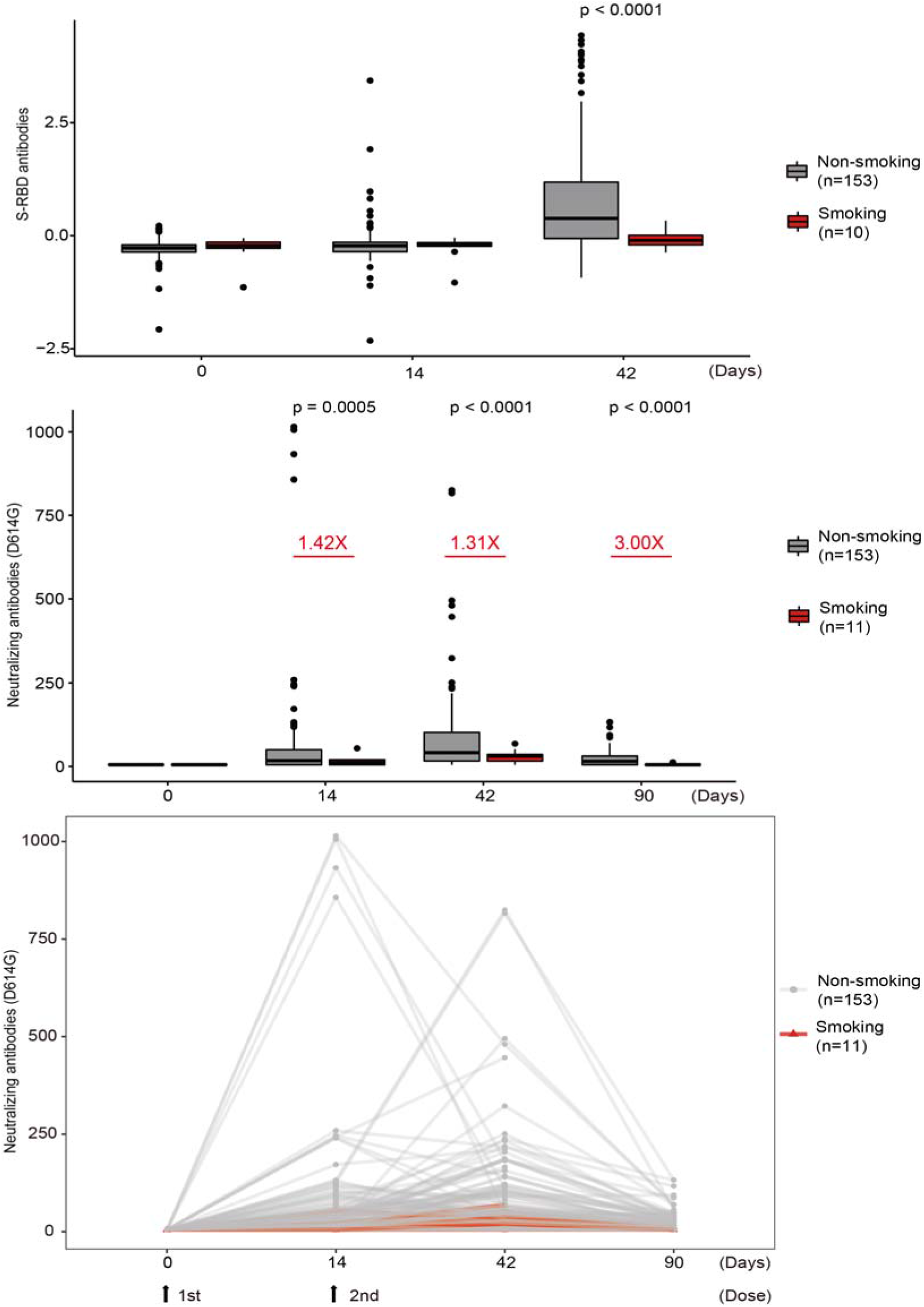
Smokers have a decreased adaptive immune response following COVID-19 vaccination compared to non-smokers. **(A)** Comparison of antibody levels to SARS-CoV-2 S-RBD between nonsmoking and smoking groups using a protein array. **(B, C)** Comparison of SARS-CoV-2 NAb levels in nonsmokers and smokers using a SARS-CoV-2 D614G pseudovirus neutralization assay represented as a (B) boxplot and (C) line chart. For (B) and (C), the y-axis represents the antibody titer that resulted in 50% pseudovirus neutralization (pNT50).

The SARS-CoV-2 Spike D614G substitution is prevalent, with data showing that it increases SARS-CoV-2 infectivity, competitive fitness, and transmission in primary human cells^5^. Therefore, we measured the levels of NAbs on days 0, 14, 42, and 90 following the first vaccine dose using a SARS-CoV-2 pseudovirus neutralization assay with the D614G substitution (Supplementary Methods). NAbs continually increased after the first and second vaccine dose, peaking on day 42 (Figure 1B and 1C). NAbs titers then significantly decreased until day 90 (Figure 1C).

Compared to nonsmokers, the NAb levels in smokers remained low throughout the period of testing. Notably, the median NAb titers in the smoking group was 1.40-, 1.32-, or 3.00-fold lower than that of nonsmoking group on day 14, 42, or 90, respectively (Figure 1B). No correlation was observed between NAbs and other factors [(i.e., age, sex, body-mass index (BMI)] (Figure S5). Altogether, our results indicate that smoking is a specific risk factor for COVID-19 breakthrough infection following vaccination. Further investigation of smoking and how it affects NAb levels in response to COVID-19 vaccination with a larger patient cohort and other COVID-19 vaccines is warranted.

## Data Availability

All data produced in the present study are available upon reasonable request to the authors.

## Acknowledgements

We thank all vaccine recipients for providing serum samples. We also thank Dr. Brianne Petritis for her critical review and editing of this manuscript. This study is supported by grants from the National Key R&D Program of China (2020YFE0202200), Beijing Municipal Education Commission (COVID-19 Emergency Response Project in Beijing Universities), Beijing Municipal Natural Science Foundation (M21003) and Beijing Municipal Science & Technology Commission (Z211102002520000), State Key Laboratory of Proteomics (SKLP-C202001, SKLP-O201904).

## References

1. Dong E, Du H, Gardner L. An interactive web-based dashboard to track COVID-19 in real time. Lancet Infect Dis 2020;20:533–4.

2. Bergwerk M, Gonen T, Lustig Y, et al. Covid-19 Breakthrough Infections in Vaccinated Health Care Workers. N Engl J Med 2021;385:1474–84.

3. Services USDoHaH. The Health Consequences of Smoking—50 Years of Progress: A Report of the Surgeon General.. Atlanta: US Department of Health and Human Services, Centers for Disease Control and Prevention, National Center for Chronic Disease Prevention and Health Promotion, Office on Smoking and Health 2014:[accessed 2016 Dec 20].

4. Lugade AA, Bogner PN, Thatcher TH, Sime PJ, Phipps RP, Thanavala Y. Cigarette smoke exposure exacerbates lung inflammation and compromises immunity to bacterial infection. J Immunol 2014;192:5226–35.

5. Hou YJ, Chiba S, Halfmann P, et al. SARS-CoV-2 D614G variant exhibits efficient replication ex vivo and transmission in vivo. Science 2020;370:1464–8.

