## Supplementary Text and Figures for "Down-regulation of SARS-CoV-2 neutralizing antibodies in vaccinated smokers"

**Supplementary Appendix**

**Table of Content**

| Supplementary Methods | page 2 |
| --- | --- |
| Supplementary Tables | page 5 |
| Supplementary Figures | page 7 |

**Supplementary Methods**

**Clinical sample collection**

Participants who received SARS-CoV-2 vaccine (Sinovac Life Sciences, Beijing, China) were recruited by the Beijing Chaoyang Hospital from December 2020 to March 2021. They received two doses of the Sinovac-CoronaVac vaccine (3 μg / 0.5 mL per dose) on days 0 and 14. Their clinical information, including underlying diseases and adverse reactions to the vaccine were collected. This study was approved by the Institutional Review Board and Medical Ethics Committee of Beijing Chaoyang Hospital (Ethical approval number: 2020-ke-491). Each recipient signed an informed consent.

**Pseudovirus neutralization assay**

The SARS-CoV-2 pseudovirus neutralization assay was performed as previously described ^1^. Briefly, serum was serially diluted three times in a 3-fold step-wise manner with a starting concentration of 1:10. The diluted serum were added to 96-well plates in duplicate and then incubated with the SARS-CoV-2 pseudovirus with a concentration of 1300 median tissue culture infective doses (TCID50/mL) at 37 ℃ for 1 hour. Monkey Vero cells were then added and incubated in a 5% CO2 environment at 37 ℃ for 24 hours. The cells were lysed and luminescent substrate was applied. The luminescence was measured, and the antibody titer resulting in 50% peudovirus neutralization (pNT50) was calculated using the Reed-Muench method.

**Detection of SARS-CoV-2 antibodies using protein array technology**

The detection of antibodies to the SARS-CoV-2 S-RBD was performed using a SARS-CoV-2 protein microarray as previously described^2,3^. Briefly, the array spotted was blocked with 5% (w/v) milk in phosphate-buffered saline with 0.05% (v/v) Tween-20 (PBST) for 10 min at room temperature. The array was then incubated with serum diluted 100-fold with PBST for 30 min at room temperature. After washing the array three times, 5 min each time, the resulting array was detected via a Cy3 Affinipure donkey anti-human IgG(H+L) antibody (Jackson ImmunoResearch, USA) and a GenePix 4300A microarray scanner (Molecular Devices, Sunnyvale, CA, USA). The median fluorescent signal intensity was extracted using GenePix Pro7 software (Molecular Devices, Sunnyvale, CA, USA).

**Statistical analysis**

The comparison of SARS-CoV-2 antibodies and NAbs between smoking and nonsmoking groups was executed using the Student’s *t*-test analysis with a significance threshold set at a p- value of 0.01. The correlation analysis between the two patient groups used a kendall correlation coefficient with a significance threshold set at a p-value of 0.01.

**References:**

1.	Nie J, Li Q, Wu J, et al. Establishment and validation of a pseudovirus neutralization assay for SARS-CoV-2. Emerg Microbes Infect 2020;9:680-6.

2.	Wang H, Wu X, Zhang X, et al. SARS-CoV-2 Proteome Microarray for Mapping COVID-19 Antibody Interactions at Amino Acid Resolution. ACS Cent Sci 2020;6:2238-49.

3.	Liang T, Cheng M, Teng F, et al. Proteome-wide epitope mapping identifies a resource of antibodies for SARS-CoV-2 detection and neutralization. Signal Transduct Target Ther 2021;6:166.

**Supplementary Table**

**Supplementary Table 1. Clinical Characteristics of the Chaoyang Cohort**

|  | **Non-smoking (N=153)** | **Smoking (N=11)** | **Overall (N=164)** | **P-value** |
| --- | --- | --- | --- | --- |
| ***Sex*** |  |  |  |  |
| Male | 28 (18.3%) | 10 (90.9%) | 38 (23.2%) |  |
| Female | 125 (81.7%) | 1 (9.1%) | 126 (76.8%) |  |
| ***Age (years)*** |  |  |  |  |
| Mean (±SD) | 33.7 (9.32) | 38.7 (11.7) | 34.0 (9.54) | 0.18 |
| Median [Min, Max] | 32.0 [20.0, 58.0] | 43.0 [23.0, 57.0] | 32.0 [20.0, 58.0] |  |
| ***BMI*** |  |  |  |  |
| <30 kg/m^2^ | 151 (98.7%) | 10 (90.9%) | 161 (98.2%) |  |
| ≥30 kg/m^2^ | 2 (1.3%) | 1 (9.1%) | 3 (1.8%) |  |

Continuous variables were expressed as mean (SD) and compared with the Student’s *t*-test. SD represents the standard deviation. Categorical variables were expressed as number of participants (%). BMI: body mass index. Obesity divides BMI into two categories^1^ , BMI < 30 kg/m^2^ as normal and BMI ≥ 30kg/m^2^ as obesity.

**Supplementary Table 2. Summary of SARS-CoV-2 pseudovirus neutralization assay results**

|  | **Non-smoking (N=153)** | **Smoking (N=11)** | **Overall (N=164)** | **P-value** |
| --- | --- | --- | --- | --- |
| ***NAb (Day 0)*** |  |  |  |  |
| Mean (SD) | 5.00 (0) | 5.00 (0) | 5.00 (0) |  |
| Median [Min, Max] | 5.00 [5.00, 5.00] | 5.00 [5.00, 5.00] | 5.00 [5.00, 5.00] |  |
| ***NAb (Day 14)*** |  |  |  |  |
| Mean (SD) | 62.2 (155) | 15.5 (14.6) | 59.1 (150) | ***0.0005*** |
| Median [Min, Max] | 17.0 [5.00, 1020] | 12.0 [5.00, 54.0] | 17.0 [5.00, 1020] |  |
| ***NAb (Day 42)*** |  |  |  |  |
| Mean (SD) | 80.5 (121) | 28.6 (19.4) | 77.0 (117) | ***<0.0001*** |
| Median [Min, Max] | 41.0 [5.00, 825] | 31.0 [5.00, 68.0] | 38.5 [5.00, 825] |  |
| ***NAb (Day 180)*** |  |  |  |  |
| Mean (SD) | 23.3 (24.5) | 6.17 (2.86) | 22.5 (24.2) | ***<0.0001*** |
| Median [Min, Max] | 15.0 [5.00, 133] | 5.00 [5.00, 12.0] | 14.0 [5.00, 133] |  |

Continuous variables were expressed as mean (SD) and compared with the Student’s *t*-test; categorical variables were expressed as number of participants (%). NAb of 5 is the lower limit of detection of the SARS-CoV-2 pseudovirus neutralization assay.

**References:**

1. Menni, Cristina et al. Vaccine side-effects and SARS-CoV-2 infection after vaccination in users of the COVID Symptom Study app in the UK: a prospective observational study. The Lancet. Infectious diseases 2021;7(21):939-949.

**Supplementary Figures**

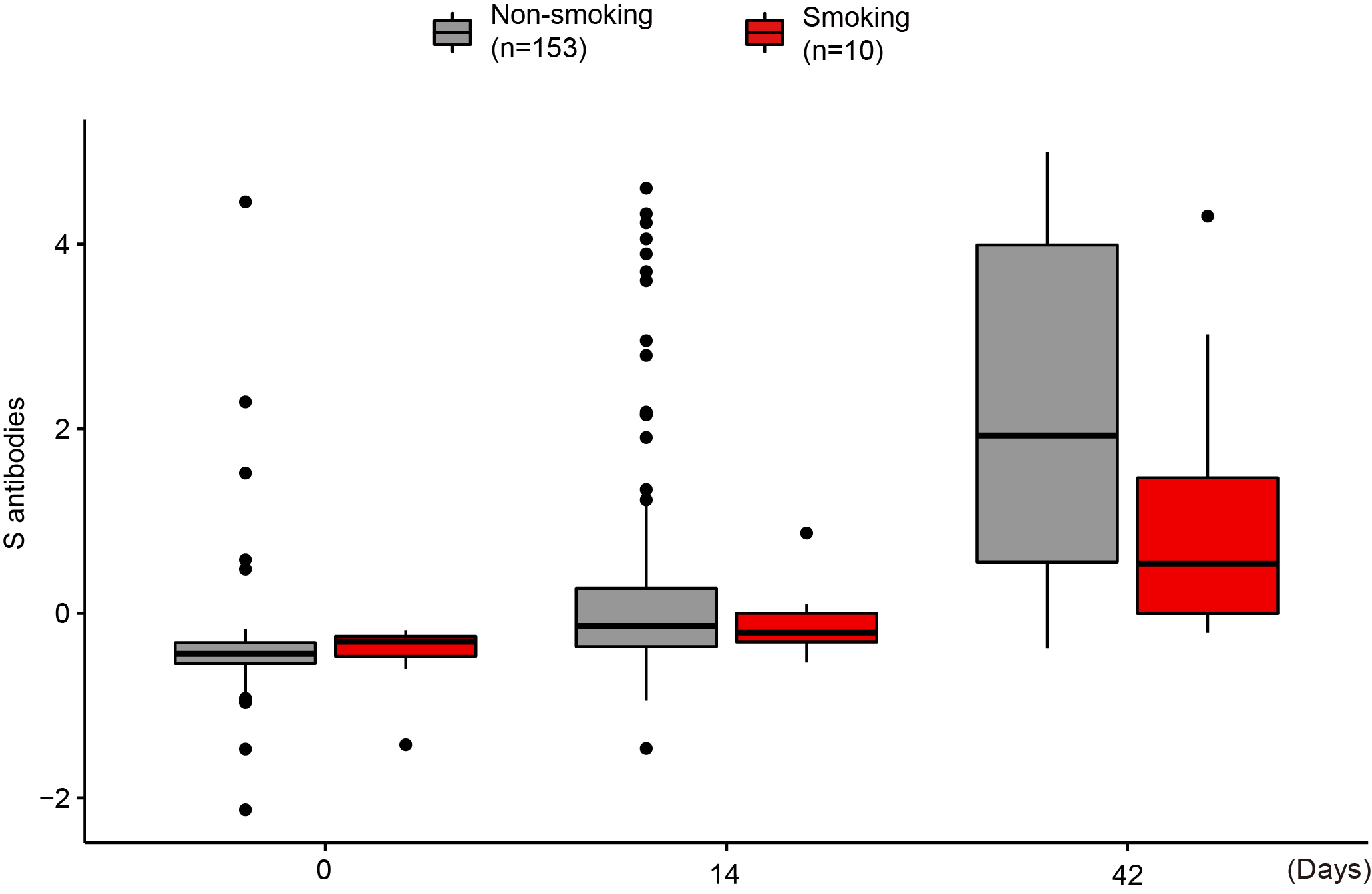

**Supplementary Figure 1. Comparison of antibody levels to the SARS-CoV-2 S-RBD after COVID-19 vaccination.**

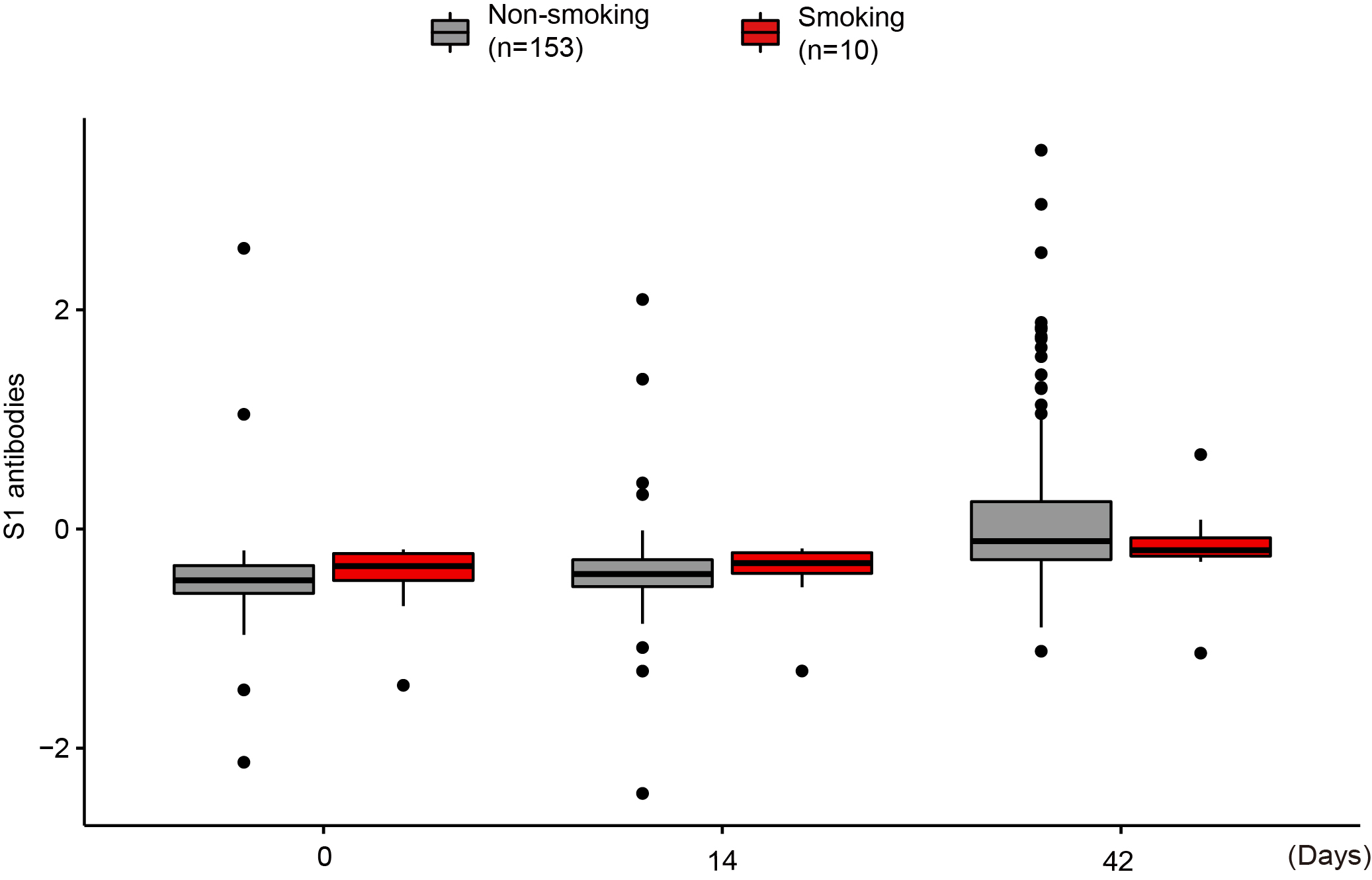

**Supplementary Figure 2. Comparison of antibody levels to the SARS-CoV-2 S1 after COVID-19 vaccination.**

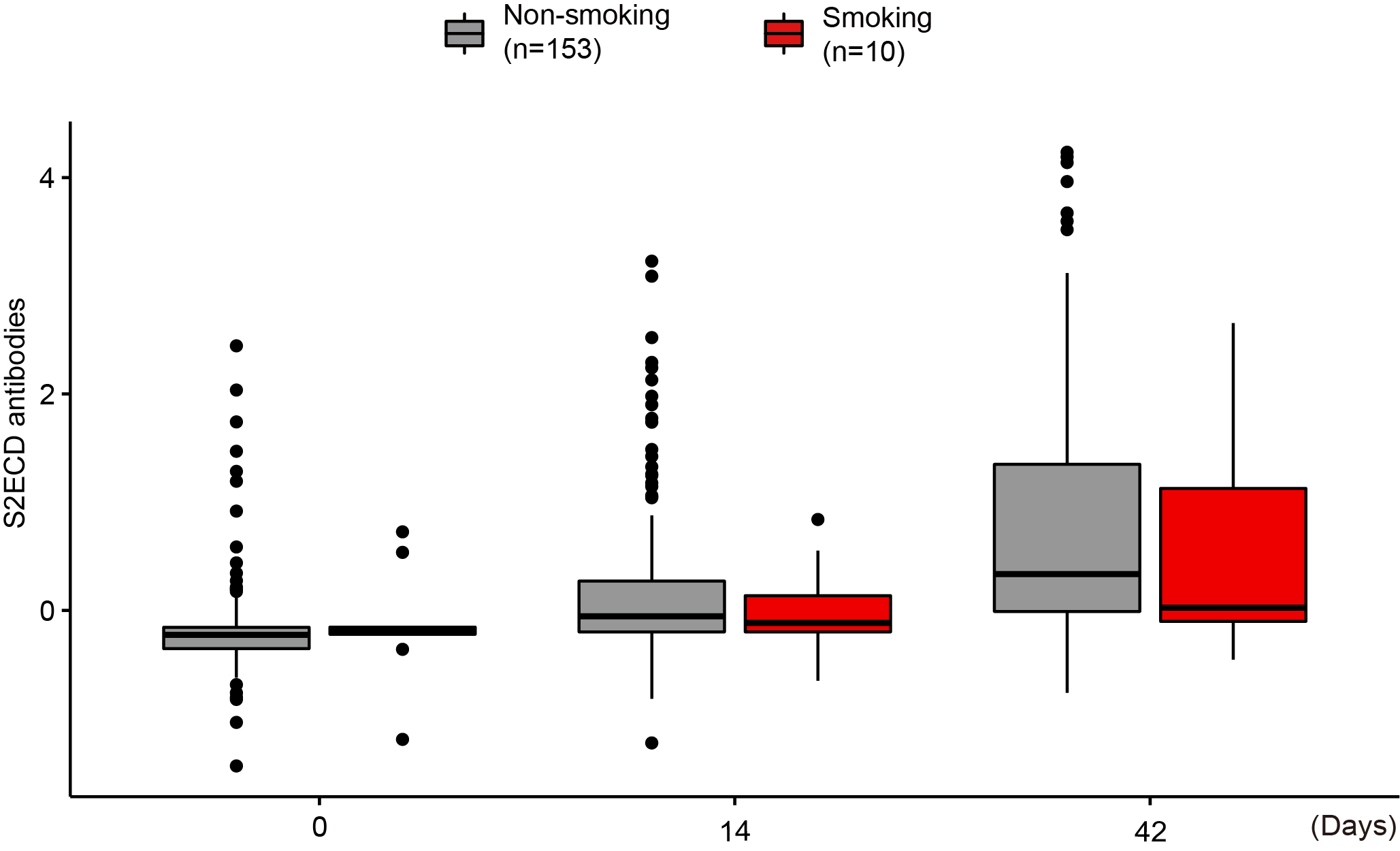

**Supplementary Figure 3. Comparison of antibody levels to the SARS-CoV-2 S2ECD after COVID-19 vaccination.**

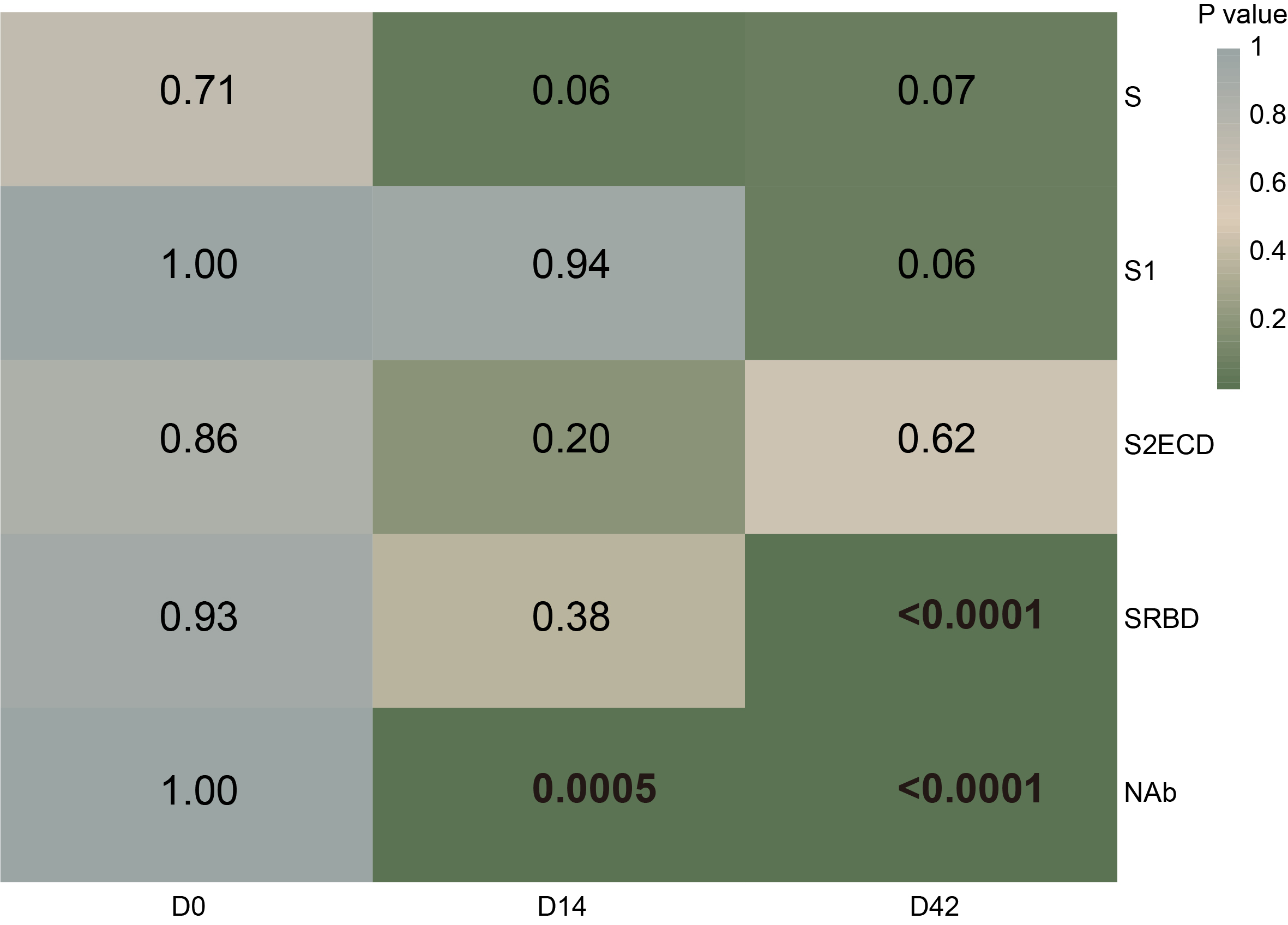

**Supplementary Figure 4. Statistical analysis of antibody levels to the SARS-CoV-2 S protein between nonsmoking and smoking groups.** “D0,” “D14,” and “D42” are 0, 14, and 42 days after the first vaccine dose, respectively. “S,” “S1,” “S2ECD,” and “RBD” represent SARS-CoV-2 antibodies targeting specific regions of the S protein. The statistical analysis was executed using the Student’s *t*-test.

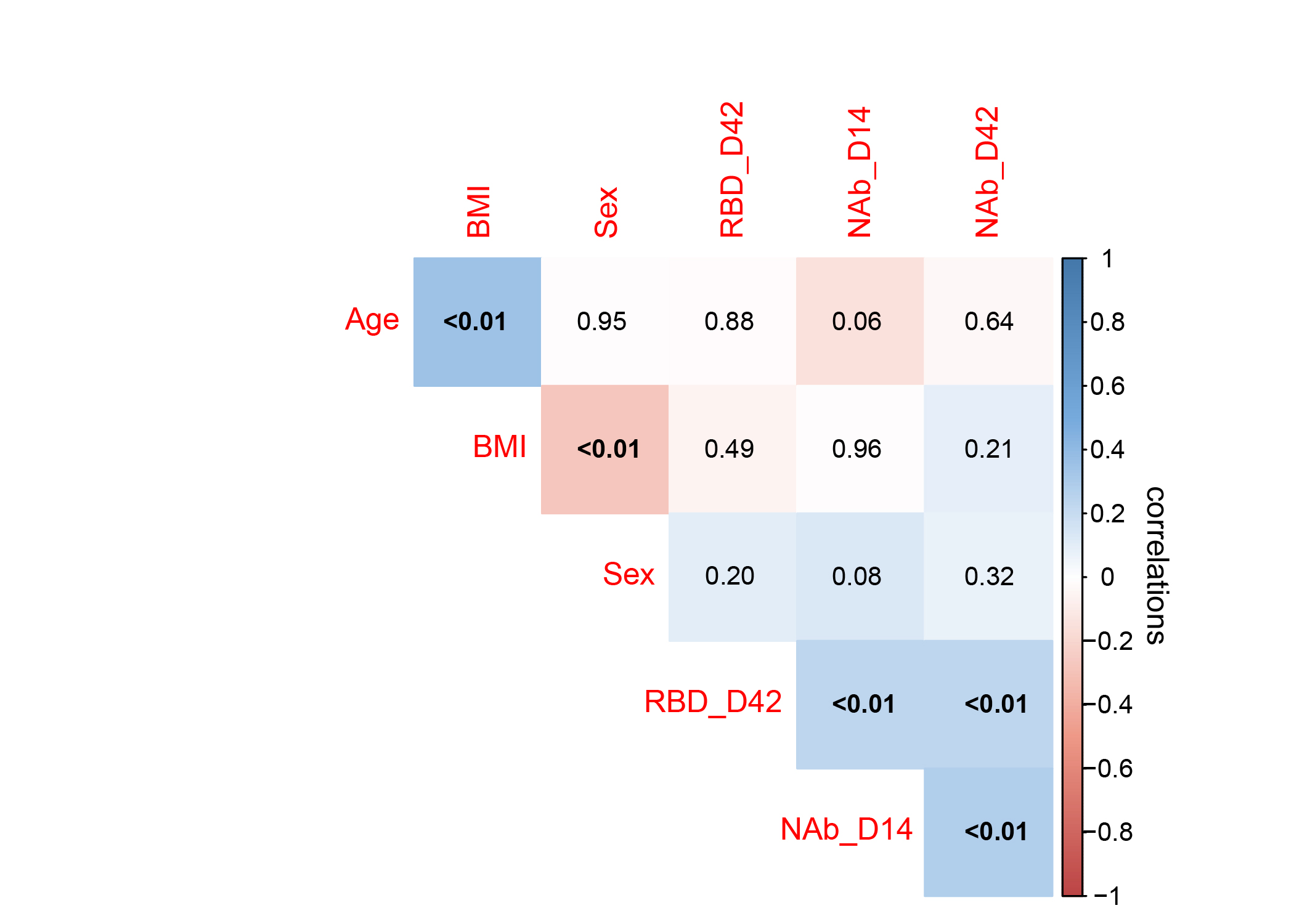

**Supplementary Figure 5. Sex and BMI are not correlated to the level of NAbs to SARS-CoV-2 S protein.** The red to blue colors represent the kendall correlation coefficient from -1 to 1, respectively. The number within each block of the heatmap represents the corresponding p-value.
